# Poultry slaughter and carcass disposal practices in Bangladesh: Piloting the use of killing cones to reduce avian influenza transmission

**DOI:** 10.1101/2025.08.30.25334525

**Authors:** Musa Baker, Kamal Ibne Amin Chowdhury, Dalia Yeasmin, Abu-Hena Mostofa Kamal, Mohammad Tauhidul Islam, Sheikh Shariful Islam, Ausraful Islam, Md. Saiful Islam

## Abstract

Mobile poultry vending and slaughtering of sick poultry have been linked to the spread of highly pathogenic avian influenza (HPAI) H5N1 in Bangladesh. However, limited data exist on associated practices and potential interventions to improve biosecurity among mobile poultry vendors. This mixed-method study was conducted in three phases across four sub-districts in Bangladesh. In phase 1, researchers conducted 416 hours of structured observation, 40 in-depth interviews with poultry vendors, and 40 informal interviews with the customers. Phase 2 involved the development and pilot testing of an intervention package, which included poultry slaughtering cones, hand sanitizers, disinfectants, and a hygiene pamphlet, with 10 vendors. Phase 3 implemented the full intervention with 20 vendors, followed by 94 hours of observation and 17 customer interviews. At baseline, vendors sourced poultry from multiple locations, kept them in small cages on rickshaw vans, and slaughtered them in open spaces, drains, or near water sources. Waste was often discarded in the environment or fed to animals. Vendors demonstrated limited hygiene knowledge and were not observed using personal protective equipment, soap, or disinfectants. Post-intervention, vendors adopted improved practices such as using killing cones, containing waste, disinfecting the slaughtering area, and cleaning hands before eating. Customers also viewed the intervention positively. The study highlights significant risks of AIV transmission through mobile poultry vending but demonstrates that low-cost, targeted interventions can enhance hygiene and biosecurity. Further research is needed to assess long-term sustainability and scalability.

## Introduction

In Bangladesh, Avian Influenza A(H5N1) virus is endemic in poultry (1). Since 2007, nine human cases of H5N1 have been reported from Bangladesh, with one death (2). All had a history of exposure to poultry or poultry products and waste before illness (3, 4). On March 15, 2011, the icddr,b urban surveillance in Kamalapur, Dhaka, identified a human case with highly pathogenic avian influenza (HPAI) H5N1 virus (4). Seven days before the illness, the patient’s mother bought seven chickens from a mobile poultry vendor. The vendor slaughtered, defeathered, and skinned the chickens, and the case’s father processed the chickens inside the case’s home while the case was present at the slaughtering site (4).

Mobile poultry vendors usually sell chickens, pigeons, geese, and ducks using a bicycle, rickshaw van, or carrying them in their hands. They often buy different species of poultry from different locations and keep them together, which contributes to the transmission and spread of the viruses. They often slaughter poultry for customers and discard waste in the environment. Disposal of slaughtering waste and carcasses in the environment are possible source of avian influenza infection in dogs, cats, and other domestic animals (5). A community-based ethnographic study in Bangladesh found that during the slaughtering of poultry, humans, animals, and birds were exposed to poultry blood, feathers, internals, and other body parts when discarded in the environment (6). Although mobile poultry vending and the slaughtering of sick poultry have been identified as sources of the HPAI H5N1 virus in Bangladesh, there is limited data available on the associated practices, including vending methods, slaughtering, and waste disposal. This study examined these practices, including poultry sourcing, selling, and slaughtering methods, waste management, and piloted an intervention to improve safe slaughtering and disposal behaviors.

## Materials and methods

### Study site

The study was conducted in four sub-districts: Mirpur, Mohammadpur, Mymensingh Sadar, and Netrokona Sadar. These locations were selected because ongoing Avian Influenza surveillance was already being conducted in their live-bird markets.

### Study design and data collection

This was a formative study where structured observation, informal interviews, and in-depth interviews were used as data collection tools. We recruited participants between 01/04/2016 and 30/04/2016. The study was conducted in three phases (Figure 1). In Phase 1, we conducted 416 hours of observation, watching each vendor from the collection of poultry to the sale of poultry for two consecutive days. We also counted the number of poultry carried by the vendors, the number of apparently sick poultry, the number of people interacting with the vendor, the customers’ direct contact with the poultry, the number of slaughtering events, including who, where, and how slaughtering was performed, and disposal of blood and other poultry waste. We also conducted 40 in-depth interviews with poultry vendors to explore poultry trading, slaughtering, and waste disposal practices, and clarified some of the practices we noticed during observation. Additionally, we conducted 40 informal interviews with poultry customers to understand the type of poultry they typically purchased from vendors, their purchasing motivations, poultry slaughtering practices, waste disposal methods, knowledge about avian influenza, and suggestions on how to minimize the spread of avian influenza to humans.

**Figure 1:**
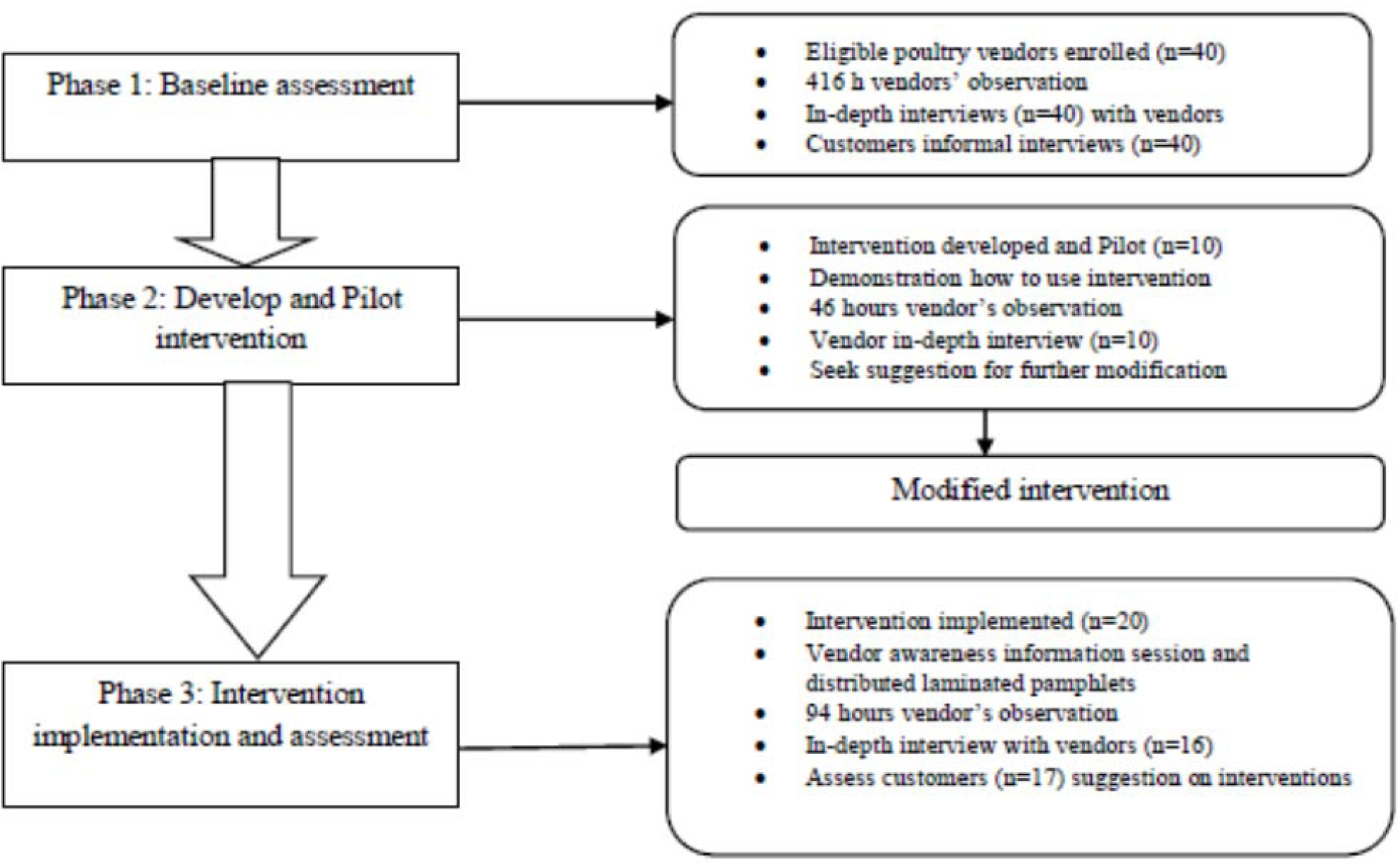
Different phases of research activities.

In phase 2, we developed and piloted an intervention package. The intervention package included a poultry slaughtering cone, which was co-designed in collaboration with the poultry vendors. We designed cones in two sizes to fit tricycles (Size-1: upper end 8” X lower end 1”) or bicycles (Size-2: upper end 6” X lower end 1”). The cones were complemented with a waste box (Length 20” X width 4” X depth 14”) with a tray (Length 20” X width 10” X depth 3”) underneath to deposit blood, where sand or sawdust was used for absorption (Figure 2). The intervention package also included hand sanitizer, soap, detergent, sodium hypochlorite, and a spray bottle, along with an informational pamphlet on personal hygiene and poultry waste disinfection illustration. Before piloting the cones, we demonstrated how to use the slaughtering cones, prepare disinfectant solutions, and wash hands using soap or hand sanitizers. The intervention was then piloted among 10 vendors: eight to those who used a rickshaw van for poultry selling and two to those who used a bicycle. After five days of the pilot, we conducted interviews with the vendors to assess the feasibility and practicality of using the cone, along with recommendations about the cone design.

**Figure 2:**
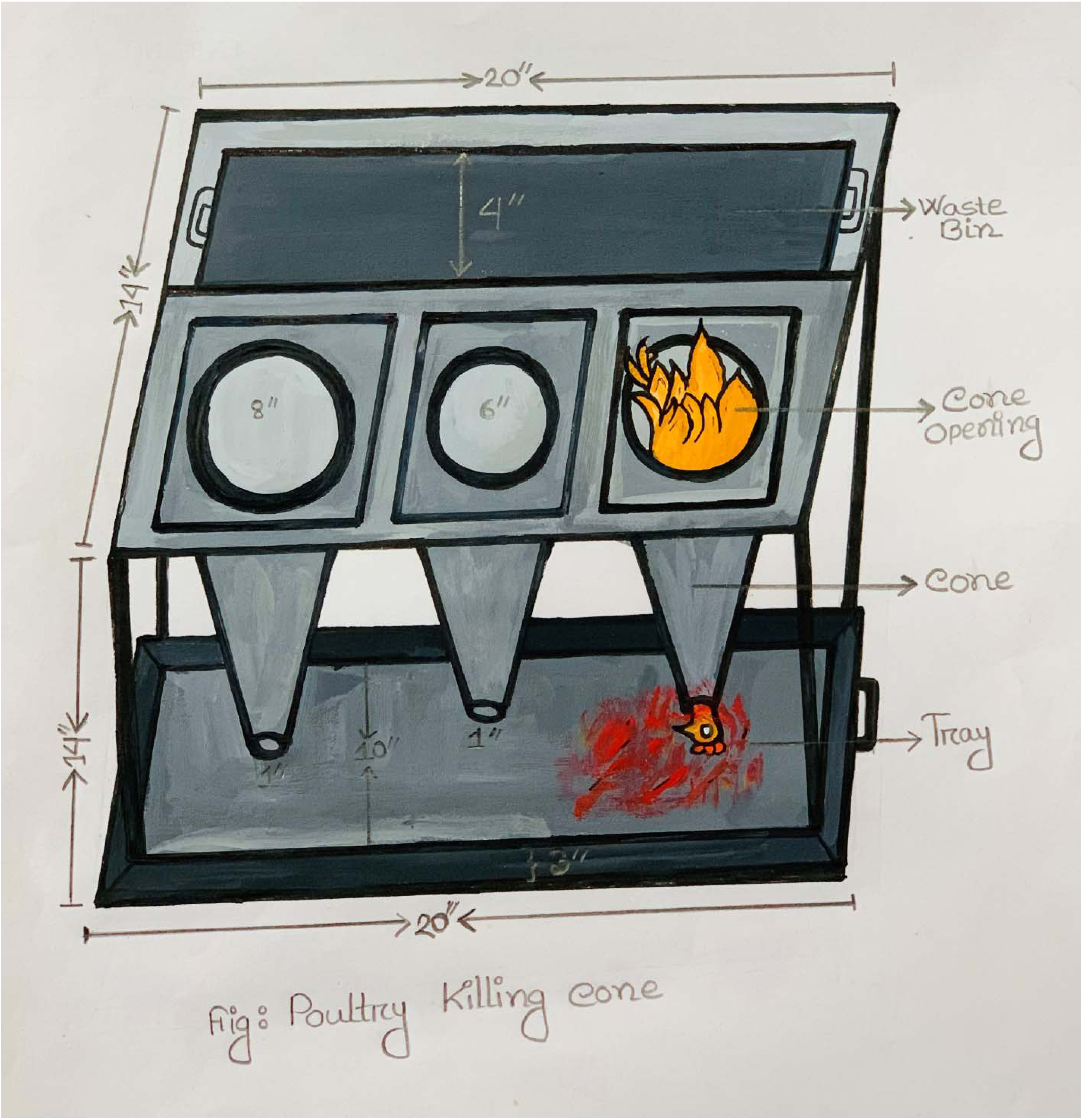
Poultry killing cone.

In the final phase, we modified the slaughtering cones based on the interview findings from the pilot phase and provided them to 20 mobile poultry vendors, each employing different approaches to selling poultry. For vendors who used bicycles, we selected and provided five slaughtering cones that were attached to the bicycles. The remaining 12 cones were distributed to vendors who used rickshaw vans to sell poultry. In areas where vendors operated from temporary selling locations, we installed three slaughtering cones in central, accessible locations to offer a safe slaughtering option for all vendors. In these locations, one vendor, selected by their peers, was designated to maintain the cone and take responsibility for slaughtering, sanitation, waste disinfection, and the supply of detergent and soap for all participating vendors. Vendors were instructed to use slaughtering cones for one month. During that period, we conducted 94 hours of unobtrusive observation and 17 informal interviews with customers to request feedback and suggestions regarding the intervention package.

### Data analysis

All quantitative data were entered and analyzed in Microsoft Excel using descriptive statistics. For qualitative data, the research team expanded field notes and transcribed all interviews verbatim in Bengali using Microsoft Word. A preliminary coding framework was developed based on the initial research questions, objectives, and was later revised through iterative reading and discussion among the investigators to incorporate emerging themes. The transcripts were then manually coded according to the final code. Coded segments were organized thematically and summarized across key domains aligned with the study objectives. Data triangulation was conducted by comparing findings across different sources, including interviews, field notes, and observations, to ensure credibility and depth of interpretation. The coding and thematic synthesis were collaboratively reviewed by multiple researchers to enhance reliability and minimize individual bias.

## Results

### Poultry trading practices

Of the 40 interviews conducted in Phase 1, 75% of vendors reported using bicycles, rickshaw vans, hands, or baskets to transport poultry for purchase and sale (Table 1).

**Table 1.** Socio-demographic characteristics of the participants.

| Characteristic | Vendors | % | Customers | % |
| --- | --- | --- | --- | --- |
| **Age (Years)** |  |  |  |  |
| Mean (SD) | 34 ±10.62 |  | 37±12.10 |  |
| **Sex** |  |  |  |  |
| Male | 40(40) | 100 | 9(40) | 22 |
| **Marital status** |  |  |  |  |
| Married | 36(40) | 90 | 40 | 100 |
| **Religion** |  |  |  |  |
| Muslims | 40(40) | 100 | 38/40 | 95 |
| **Educational status** |  |  |  |  |
| No formal education | 8(40) | 20 | 8(40) | 20 |
| Primary education | 16(40) | 40 | 9(40) | 40 |
| Lower secondary education | 14(40) | 35 | 11(40) | 35 |
| Higher secondary or above | 2(40) | 5 | 12(40) | 5 |
| **Monthly income (BDT)** |  |  |  |  |
| Mean | 16025±5245.20 |  | 29712± 28,001 |  |
| **Alternative occupation** |  |  |  |  |
| Poultry trading | 19(40) | 48 | 0(40) | 0 |
| Agriculture | 15(40) | 38 | 0(40) | 0 |
| Others | 6(40) | 15 | 0(40) | 0 |
| **Poultry business involved persons** |  |  |  |  |
| Family member | 18(40) | 45 | 38(40) | 95 |
| Relatives | 19(40) | 48 | 2(40) | 5 |
| Friends | 3(40) | 7.5 | 0 | 0 |

Vendors reported purchasing 20-30 poultry daily from wholesale or village markets. In addition to poultry, three participants reported selling water birds, including cormorants, egrets, herons, and migratory birds. Unsold poultry was reportedly kept in a cage in their living room or separated in another cage at night. Vendors reported selling both healthy and sick poultry, specifically reducing the price to sell sick poultry. Vendors reported that they attempted to sell sick poultry first and kept the poultry fresh by spraying them with water. During observation, we found 40 vendors carried 2,411 poultry; 19% appeared sick, and one died (Table 2).

**Table 2.**
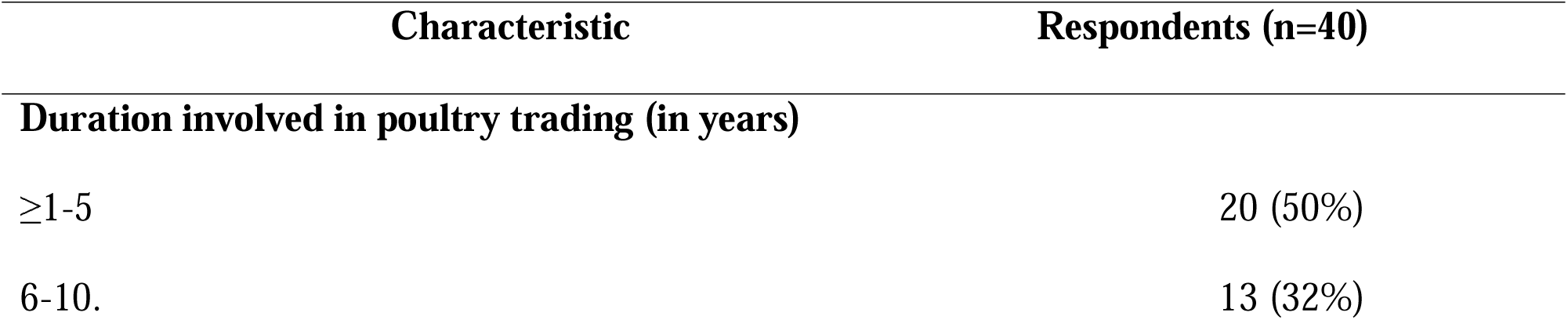
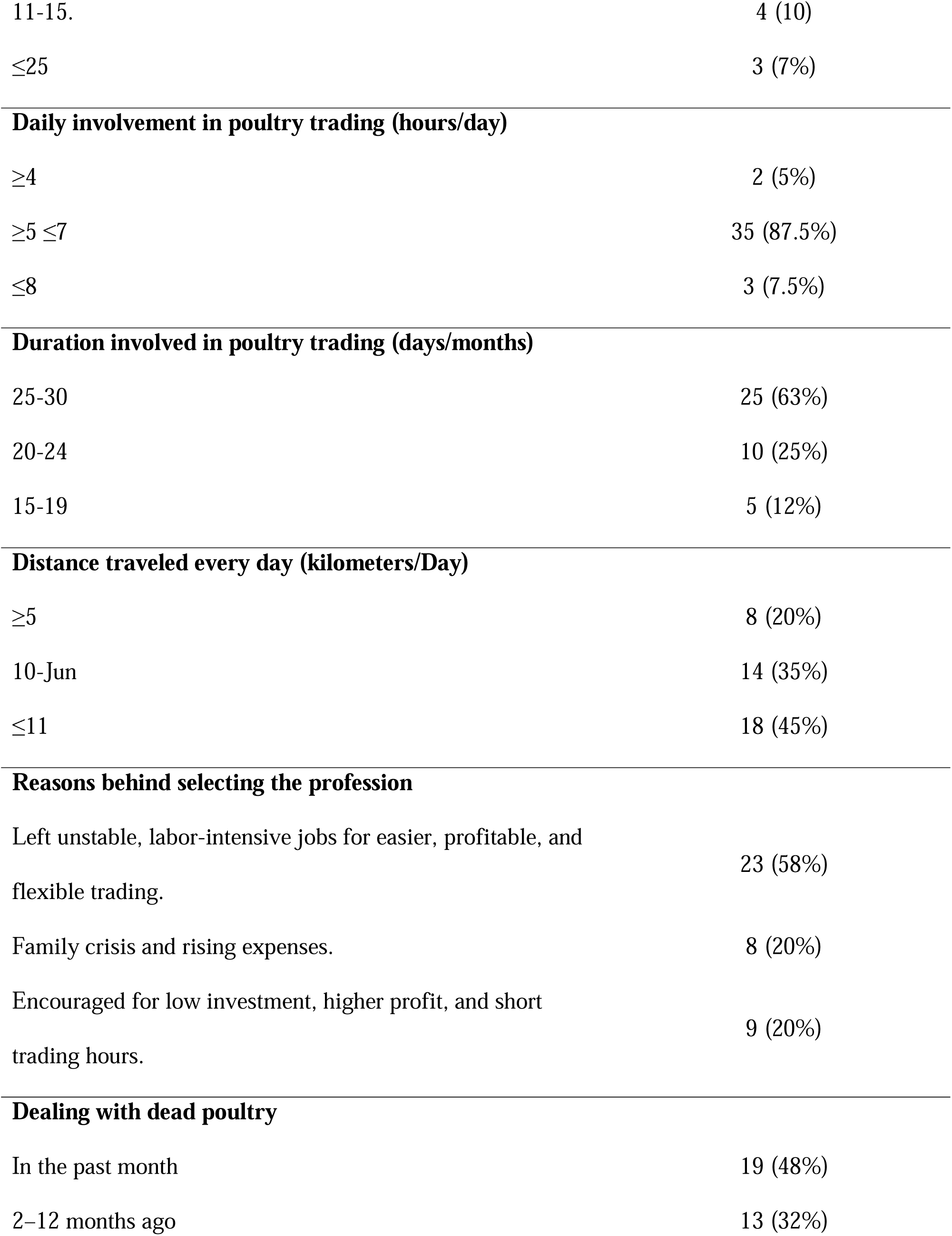
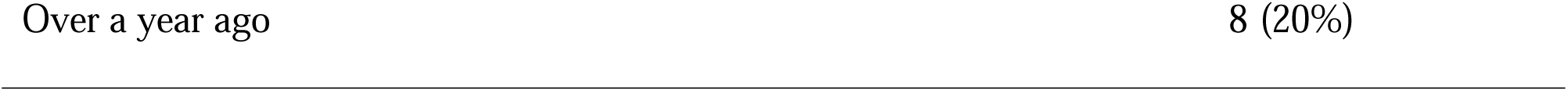
Occupational History of Mobile Poultry Vendors.

### Poultry slaughtering and waste disposal practices

Eighty percent of vendors reported slaughtering both healthy and sick poultry. Slaughtering was commonly carried out in customers’ yards, bathrooms, in a plastic jar, along roads, in drains, or near water sources, which was also confirmed through observation. We observed the slaughtering of 1,959 poultry by vendors, and personal protective equipment (PPE) was not used in any instance. In 442 cases (23%), vendors sought assistance from consumers during the slaughtering process. They performed slaughtering in a plastic jar (1,139 cases; 58%), directly on the road (415 cases; 21%), in the roadside drain (214 cases; 11%), and on newspapers (39 cases; 2 %). Seventy percent of vendors reported discarding slaughtering waste directly into the environment. The remaining 30% reported storing waste in plastic drums, buckets, polythene bags, or sacks, and attempting to sell it to fisheries or low-income individuals who consume poultry skins, legs, and internal organs. Most vendors indicated that waste or dead poultry was discarded on roads, agricultural land, drains, open areas, bushes, ditches, ponds, or rivers. We observed vendors frequently disposing of slaughtering waste by feeding it to crows and stray dogs.

### Hygiene practice and use of personal protective equipment

Nearly 90% of vendors reported being unaware of proper hygiene practices and had never used PPE, such as face masks, aprons, gloves, or head covers while handling poultry. Almost all vendors (38 out of 40) reported not washing their hands after handling poultry. Vendors reported washing their rickshaw vans or bicycles once per week, while cages were cleaned either daily or once every two weeks, depending on the location. Cages were washed in rivers, ponds, ditches, or in their homes. Slaughtering equipment was typically wiped with a rag or rinsed with water; however, most vendors reported cleaning the slaughter drum with detergent. All participants stated that they washed their rags and clothes with soap daily. We observed vendors repeatedly wipe their hands after slaughtering, using a handloom towel (Gamsa), a handkerchief, or a piece of cloth. We also observed vendors taking snacks while trading without cleaning their hands.

### Risk perception about avian influenza

Nearly half of the vendors were unaware of the term HPAI; however, they were able to describe the clinical symptoms observed in poultry. One-third of vendors recognized that poultry diseases are contagious and can be transmitted. The majority of vendors (78%) reported not knowing how diseases could spread from poultry to other animals or humans, nor were they aware of preventive measures. Additionally, most vendors were unaware of their potential role in disease transmission and prevention, including the importance of safe slaughtering practices.

### Phase 2: Intervention development and piloting

The vendors reported that the cones and waste boxes helped keep their vans clean and free of odor. All participants stated that the pamphlet served as a useful reminder for when to disinfect their hands with sanitizer and how to disinfect and discard waste properly. Bicycle users, however, reported mixed experiences with the use of slaughtering cones. Bicycle users reported that the killing cone was noisy, heavy, and reduced balance and speed. Also added that carrying hand sanitizer and disinfectant separately was inconvenient due to a lack of a carrier.

We observed, cleaning the cone was difficult, as dried blood often stuck to its walls.

### Phase 3: Implementation and assessment

#### Acceptability of the intervention

Vendors reported that the slaughtering cone and waste box helped keep their tricycles clean and odor-free. They expressed satisfaction with the cone, describing it as a simple tool that reduced visible blood and poultry waste in the environment. Following the intervention, vendors acknowledged that hand sanitizer and waste disinfectants could help prevent disease transmission. Consistent with findings from the pilot phase, vendors using bicycles reported challenges with the slaughtering cone; one vendor described issues such as jerking, noise, heaviness, reduced speed, and difficulties with balance. Two participants recommended using lighter materials for cones. During participant observation, a total of 294 poultry were slaughtered. Of these, vendors used the killing cone in 232 cases (79%), while slaughtering was conducted on the roadside in 33 cases (11%), beside roadside drains in 7 cases (2%), and in other places in 22 cases (7%). Waste materials such as blood, feathers, and intestines were stored in the killing cone’s attached waste boxes in 228 instances (97% of cone uses). Vendors were observed consuming snacks, including biscuits, water, tea, and betel leaf. In 75% of cases, vendors cleaned or rubbed their hands with hand sanitizer before eating. Along with, vendors sprayed the Sodium Hypochlorite (NaOCl) on wastages 39 times after each episode, and 75% of vendors discarded the final wastages in the city corporation-provided dustbin. Informal interviews with customers also revealed positive perceptions of the intervention. Most customers (88%) recognized the cone’s role in safe waste disposal, and 53% considered it a useful tool overall. Additional benefits identified by customers included access to freshly slaughtered poultry (29%), convenience during the slaughtering process (24%), and protection from bacterial contamination (18%) from previous slaughtering wastes. Customers also noted that the cone helped maintain a clean environment and could contribute to disease prevention.

## Discussion

Baseline findings indicated that mobile poultry vending practices create a high-risk environment for the spillover of HPAI H5N1 from poultry to other poultry, poultry to animals such as dogs or crows, and poultry to humans. This risk is driven by factors such as multiple poultry sourcing, mixing them in a cage, unsafe selling and storage practices, inadequate cleaning procedures, poor waste disposal, and the handling of sick or dead birds. The intervention comprising a poultry slaughtering cone, waste storage system, and an educational pamphlet on hand hygiene and waste disposal likely contributed to improvements in safe slaughtering practices, waste management, and risk perception related to avian influenza viruses. The slaughtering cone intervention was well accepted by both vendors and consumers. Further research is recommended to evaluate the long-term effectiveness and scalability of the slaughtering cone approach. Since the introduction of HPAI H5N1, a total of 585 outbreaks have been reported across 54 of the country’s 64 districts, positioning Bangladesh among the nations with the highest number of documented cases globally (7). A Bangladeshi study found influenza in 18% of severe acute respiratory infection cases and noted that 59% of related deaths occurred post-discharge (8). Our study found that vendors sourced multiple poultry species from various locations and housed them together in single cages with poor ventilation and without regular cleaning or disinfection, which increases the risk of AIV transmission among poultry (9). Besides, the findings of slaughtering poultry along roads, in drains, or near water sources increase the risk of transmission to other animals such as dogs and crows. These behaviors likely contribute to the recurrent outbreaks of HPAI H5N1 observed in crows in Bangladesh (10).

Our findings also highlight critical risks of animal-to-human transmission. Mobile poultry vendors pose a potential risk of AIV transmission through the transportation of live poultry from markets to residential neighborhoods, the slaughtering of sick birds, and the lack of facial protection during handling (11). A companion study (unpublished) detected H5 virus in 2% of swabs from healthy poultry and found influenza A viruses present year-round. Vendors transported poultry over long distances, indicating a potential role in spreading zoonotic influenza. During the slaughtering and bleeding process, flapping poultry can generate droplets containing viral particles, which may settle on surrounding surfaces within one meter of the source (12). Mobile poultry vendors’ daily movements with potentially infected poultry, contaminated products, and equipment could increase the risk of exposures to AIVs (13).

Limited knowledge of AIV transmission pathways, combined with the lack of handwashing after slaughtering poultry and before eating, increases the risk of AIV transmission among poultry vendors. Similar exposures have been linked to H5N1 infections among poultry market workers in Bangladesh (14, 15). International guidelines discourage slaughtering poultry outside of designated slaughter facilities or on farms; however, low-income populations in developing countries are unlikely to adopt this practice, as mobile poultry vending practices are deeply rooted in cultural customs, rituals, and beliefs (16, 17). To enhance biosecurity, minimize the risk of product contamination, and reduce vendor and customer exposure, our intervention was likely acceptable and practicable. The intervention not only increased participants’ knowledge about disease transmission pathways but also translated that knowledge into improved practices. Enhanced safe slaughtering behaviors and hand washing attracted more customers and increased customer satisfaction, indicating the potential scalability of the intervention. The findings underscore the need to scale up and evaluate this intervention that live bird mobile poultry vendors and market workers would be willing and able to implement. Slaughtering chickens in a containment vessel (killing cone), with blood collected in a bucket and waste stored in a covered container, is likely to reduce the risk of virus transmission not only through airborne and droplet routes but also via environmental contamination (18). This approach aligns with the recommendations provided by the World Health Organization (WHO) (19) and plays an important role in mitigating the transmission of diseases. A study recommended prioritizing interventions to enhance the safety of the slaughtering environment to reduce disease transmission, and to enhance sanitation and waste disposal facilities (20). Vendors readily adopted the slaughtering cones and expressed satisfaction with their design. By using a co-design approach, we ensured that the intervention and associated behavior change messages were contextually appropriate, feasible, and accurate, ultimately aiming to improve the health environment and reduce disease burden. The intervention increased awareness of personal hygiene practices, which translated into greater use of hand sanitizer and more consistent disinfection of waste. In a study conducted in 12 live bird markets across four regions of Bangladesh, workers who frequently handled poultry, cleaned faeces, and food containers without washing their hands had a 7·6 times higher risk of H5N1 infection compared to those who practiced hand hygiene (21). Our study revealed a lack of hand hygiene among mobile poultry vendors, consistent with the findings from a study on quail rearing practices in Bangladesh, where workers reported no personal hygiene practices and were observed wiping blood from their hands with towels or clothing after slaughtering (22). In our study, increased handwashing among vendors suggests a potential reduction in infection risk. Supporting this, previous research has demonstrated that alcohol-based hand sanitizers containing 62–70% alcohol rapidly inactivate HPAI H5N1, further underscoring their value in disease prevention (23). Slaughterhouses and poultry waste act as reservoirs for pathogens capable of infecting both humans and animals. Thus, safe and effective waste disposal methods are critical for mitigating disease transmission(24, 25). Proper treatment of poultry waste not only reduces infection risks but also contributes to environmental sustainability through the generation of valuable by-products (26). In our study, vendors sprayed 10% sodium hypochlorite inside the waste storage containers after each slaughtering occasion and disinfected the waste bins at the end of each trading day. Evidence indicates that both chlorine and non-chlorine oxidizing agents are effective in reducing HPAI H5N1 contamination in poultry farm environments (27). Continued and expanded implementation of slaughter cones, disinfection protocols, and hygiene materials among mobile poultry vendors could meaningfully reduce the risk of HPAI transmission in Bangladesh.

Our study has several limitations. First, it was conducted in only four sub-districts, and the duration of observational activities was limited. However, our findings are consistent with studies conducted in live bird markets in other regions of Bangladesh. Second, as researchers observed multiple customers simultaneously, some activities may have been missed, potentially leading to an underestimation of the true frequency of trading practices. Third, we did not conduct testing for the H5N1 virus, which limited our ability to directly assess transmission risk. Effective biosecurity is required to safeguard poultry health, minimize disease spread, and reduce risks to poultry workers and the surrounding environment (28). Similarly, the effectiveness of the intervention in improving vendor slaughtering safety was not formally evaluated. Poultry vendors may also be exposed to various strains of avian influenza due to handling birds without personal protective equipment and sourcing poultry from multiple locations. Poultry vendors and other human activities have a crucial role in disease dissemination, with workers and visitors potentially serving as carriers of infectious agents on poultry farms (29). Finally, the study did not assess the long-term feasibility of the intervention, nor did it include follow-up monitoring to evaluate the sustainability of behavioral changes over time.

## Conclusion

The baseline findings highlight critical risks at the human–animal–environment interface. The study’s findings demonstrate that the utilization of slaughtering cones raised awareness about the risk of HPAI transmission among vendors and improved safe slaughtering practices and waste disposal. To ensure sustained reductions in HPAI transmission, future initiatives should integrate individual and community-level killing cone interventions, awareness campaigns, and regulatory action, while also implementing longer-term strategies to assess the scalability and effectiveness of the intervention. Additionally, incentivizing safe and environmentally responsible slaughtering cone operations can minimize environmental contamination.

## Contributors

MSI and AFI conceptualized the study. MSI and AFI developed the methodology. MSI, AFI, and MB lead the investigation. Resources were provided by MSI, AFI, MB, KIA, DY, and AMK. Formal analysis was conducted by MB, DY, AMK, and KIA. The original draft was prepared by MB, DY, AMK, and KIA. MSI, MB, and AFI provided supervision. Funding was acquired by MSI and AFI. Visualization was done by MB, AMK, DY, and MTI. Project administration was managed by MB, MTI, and SSI. All authors contributed to reviewing and editing the manuscript. All authors had full access to the data, critically reviewed the manuscript for important intellectual content, and approved the final version. MSI, AFI, and MB had the final responsibility for the decision to submit for publication.

## Data sharing statement

Following the data policies of the contributing institutions, the primary data supporting this study cannot be made publicly available to protect intellectual property rights. However, data may be made available upon reasonable request and subject to approval by the institutional Data Access Committees of the contributing institutes.

## Conflicts of Interest

We obtained written informed consent from study participants. The study protocol was reviewed and approved by the Institutional Review Boards at icddr,b. The authors have no potential conflicts of interest to declare. The authors disclosed no conflicts of interest, and the funders were not involved in the study’s design, data collection, analysis, decision to publish, or preparation of the manuscript.

## Funding statement

The research was financially supported by the Centers for Disease Control and Prevention (CDC), which had no role in study design, data collection, analysis, or publication decisions.

## Data Availability

Data cannot be shared publicly because of ethical restrictions related to participant confidentiality and informed consent. Data are available from the icddr,b Institutional Data Access/Ethics Committee (contact via email) for researchers who meet the criteria for access to confidential data.

## Acknowledgments

The authors are grateful to the field team members and study participants from the Dhaka, Mymensingh, and Netrokona districts for their valuable contributions to this project. Sincerely, thank Fiza Feha for preparing the poultry killing cone illustration used in this manuscript. icddr,b gratefully acknowledges the Government of Bangladesh and the Government of Canada for their generous core/unrestricted support. Artificial intelligence tools were used to enhance the readability and English language expression of this manuscript.

## Notes

### Competing Interest Statement

The authors have declared no competing interest.

### Funding Statement

The author(s) received no specific funding for this work.

### Author Declarations

We obtained written informed consent from study participants. The study protocol was reviewed and approved by the Institutional Review Boards at icddr,b.

